# EMSNet: A neural network model with a self-attention mechanism for prehospital prediction of care needs

**DOI:** 10.1101/2020.05.27.20113290

**Authors:** Joo Jeong, Yu Jin Kim, Dae Kon Kim, Tackeun Kim, Joonghee Kim

## Abstract

**Background:** An artificial intelligence (AI) system capable of predicting patient needs in the prehospital phase would be instrumental. We sought to develop a neural network (NN) model capable of predicting various care needs at initial contact by emergency medical service (EMS) using multimodal input data.

**Methods:** We used EMS records of a single emergency department (ED). We implemented two attention-based NN model (I and P) differing only by how they use contextual information. The models predict multiple events, including hospital admission, endotracheal intubation, mechanical ventilation, vasopressor infusion, cardiac catheterization, surgery, intensive care unit (ICU) admission, and cardiac arrest. The input features include both unstructured data (chief complaints, injury summary, past medical history, history of present illness) and structured data (age, sex, pupil status and initial vital signs, level of consciousness, and O2 saturation on pulse oximetry). We applied multi-task learning for training. We evaluated the relative performance of the models compared with a human expert, an emergency physician with 10-year experience as an EMS medical director.

**Results:** The study population included 42,073 cases. The receiver operating characteristics (ROC) area under the curve (AUC) values of the models I and P ranged from 0.793 to 0.929 and 0.812 to 0.934, respectively. The precision-recall (PR) AUC values ranged from 0.149 to 0.673 and 0.156 to 0.683, respectively. With decision thresholds set to achieve equivalent recall levels, our AI models achieved precision levels not significantly different from those of a human expert except in prediction of mechanical ventilation and ICU admission, where the models achieved superior performance (p=0.030 [model I] and p=0.015 [model P], respectively).

**Conclusions:** AI models using multimodal input data can predict medical resource requirements at initial contact by EMS with high accuracies.

## 1. Introduction

Accurate prediction of patients’ needs is critical in prehospital care because the type of hospitals and subsequent cares are dependent on it^1,2^. Erroneous ones may lead to inefficient care delivery after transport and, ultimately, poor outcomes. However, the task is not easy. It requires an accurate assessment of both the current condition and the possible future events of a patient, which needs significant knowledge and experience in medicine. Direct medical control by emergency medical services (EMS) directors has been providing essential clues in challenging cases^3-6^. However, it means an increased workload for the directors, and maintaining such support 24/7 is sometimes impossible. Therefore, artificial intelligence (AI) system capable of doing the task with similar performance as a human expert will be a useful resource.

Achieving human expert-level performance requires being able to process unstructured natural language data efficiently as well as structured tabular data. Also, the system should be able to focus on relevant information because natural language data have diverse information.

In this study, we present our self-attention based AI system, EMSNet. It encodes natural language data with contextual information and applies a self-attention mechanism to focus on relevant information. Using the encoded representation and tabular form data, it jointly predicts various hospital resource requirements of a patient. We trained the system using multi-task learning (MTL) methods, and the system achieved human expert-level performance in our experiment.

## 2. Materials and methods

### 2.1. Study setting and population

The study is a single-center observational study utilizing EMS records of the patients who visited the emergency department (ED) using public EMS from 2011 to 2015. The study facility is a tertiary academic hospital located in South Korea with an annual ED visits greater than 80,000 patients a year. We excluded out-of-hospital cardiac arrest (OHCA), dead on arrival (DOA), and transferred-out cases. Recurrent visits were treated as independent cases. The institutional review boards of the study site approved the study and provided a waiver of informed consent.

### 2.2. AI tasks

We applied multi-task learning with one main task and five auxiliary tasks^7^. The main task is a multiple binary prediction problem with its targets include hospital admission, endotracheal intubation, mechanical ventilation, vasopressor infusion, cardiac catheterization, surgery, ICU admission, and cardiac arrest within 24 hours of ED arrival. The auxiliary tasks include the prediction of primary ED diagnosis and ED disposition using the final output of the shared portion of the network (auxiliary task group 1) and performing the main task and the two auxiliary tasks using the intermediate output of the shared network (auxiliary task group 2). The detailed description of the tasks is in supplementary table 1.

### 2.3. Datasets

We used EMS records of the study facility from 2011 to 2015. We used features obtainable at the initial encounter by EMS to make the AI system to be able to predict the resource needs and thus the destination of transport as soon as possible. The features were age, sex, chief complaints (CC), injury summary (if injury-related), past medical (and surgical) history (PMH), history of present illness (HPI), pupil status (size and reflex), systolic blood pressure (SBP, mmHg), diastolic blood pressure (DBP, mmHg), pulse rate (PR, beats per minute), respiratory rate (RR, breaths per minute) and body temperature (BT, measured in Celsius), level of consciousness (AVPU: Alert, Verbal, Pain and Unresponsive), initial O_2_ saturation (SpO_2_ on pulse oximetry, %). Free-text data (CC, injury summary, PMH, HPI) were cleaned, lower-cased (for alphabet words), and space-corrected. The target variables of the primary and auxiliary tasks were extracted from the electronic health record (EHR) database.

The dataset was randomly split into training, validation, and test sets with the ratio of 6:2:2. The training dataset was used to develop preprocessing pipelines and to train models. The validation dataset was used to evaluate candidate models and their hyperparameters. The test dataset was used to measure the performance of the final models.

#### 2.4.1. AI system: preprocessing pipelines

The preprocessing pipelines for the multimodal input data are developed using the training dataset and include the following procedures: 1) The NLP pipeline tokenizes and index natural language data using a Korean natural language processing (NLP) tool, soynlp^8^; 2) The pipeline then embeds the tokens using FastText algorithm (implemented in Gensim library) and inhouse-corpus based mainly on Korean Wikipedia and the HPI part of the training dataset^9^. 3) The non-NLP pipeline does the other common preprocessing procedures where categorical features are one-hot encoded, and numerical features are standardized by removing their means and scaling to their unit variances. Missing values are imputed by the means or the modes as appropriate with adding missing indicators to the datasets.

#### 2.4.2. AI system: NN architecture

We assumed a typical healthcare provider would read a medical note in the following sequence, which our EMSNet architecture tries to mimic: First, the reader will briefly look at the contextual information, such as chief complaints, demographics, and underlying conditions. Then the reader will read the whole HPI using the contextual information. Lastly, the reader will interpret various measurements, such as vital signs, exam findings, and test results. If the reader wants to predict a specific health outcome event, he or she can go back to the note and reread the data, focusing on specific parts of the text relevant to the outcome event.

Figure 1 visualizes the computational graph of EMSNet, which follows a similar process. Briefly, the demographic information (age and sex) and other contextual information (CC, injury summary, and PMH) are concatenated and then transformed by two consecutive fully-connected (FC) layers to output a latent contextual vector ***c***. This vector is fed into a *l*-layer bidirectional gated recurrent unit (GRU) network with a self-attention mechanism where HPI is fused with the contextual information and turned into a sentence embedding. In this process, the contextual information is used in two different ways, either by being overwritten on the initial hidden states of GRUs (model I) or being concatenated with each word embedding vectors of HPI (model P). Then the *d_h_* -dimensional hidden state vectors *h*s of the GRUs are concatenated at each timestep forming an output matrix *H* with *n*-by-2*ld_h_* shape (2 for bidirectional), which is then processed by an attention mechanism proposed by Lin et al.^10^ where attention weights ***a*** for *H* is derived from the same *H* using the following formula:

**Fig. 1.**
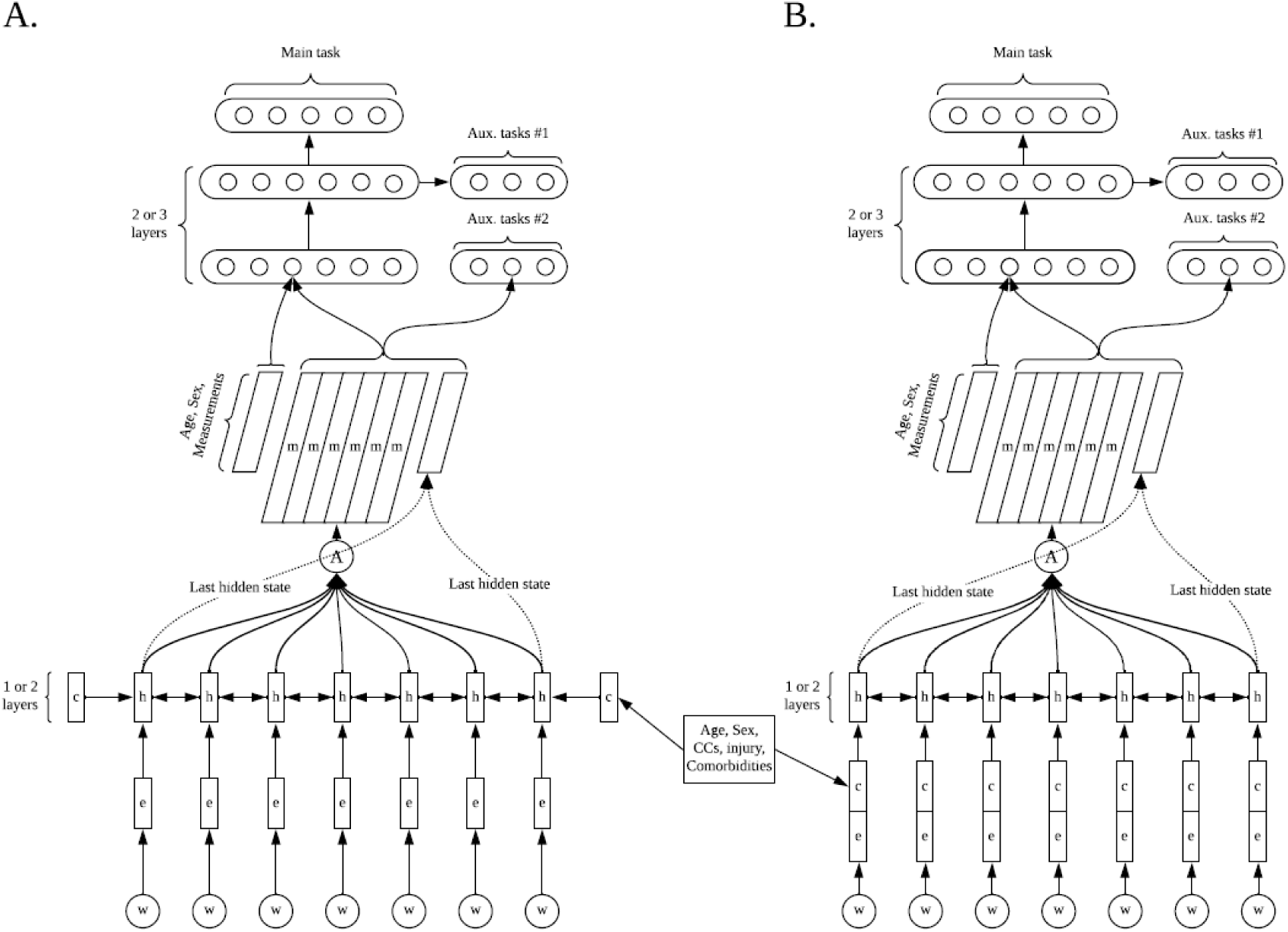
Network schematics of EMSNet. The model I (A) and P (B) are differed by where the contextual information is incorporated into the computational graph; CC, chief complaint

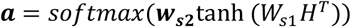

Here *W_s_*_1_ is a weight matrix with a shape of *d_a_*-by-2*ld_h_* and ***w_s_*_2_** is a vector of size *d_a_* where *d_a_* is a hyperparameter we can optimize. With ***a***, the hidden state matrix *H* is summed up to obtain a representational vector ***m*** of HPI. To obtain multiple representations from a sentence, we extend ***w_s_*_2_** into a *r*-by-*d_a_* matrix ***W_s_*_2_** resulting in the attention weight vector ***a*** becoming an attention matrix *A*. Finally, we obtain a sentence embedding matrix of HPI, *M* by multiplying *A* and *H*, which is then flattened and concatenated with the final hidden state vectors of the GRU network and the standardized measurements (vital signs, consciousness, pupillary status, and SpO_2_) and fed into a sequence of FC layers. Also, there are six task-specific networks with two FC layers, one for the main task and five for the auxiliary tasks.

#### 2.4.3. AI system: loss function

The loss function has two terms. The first one is the weighted sum of cross-entropy losses from the task-specific networks with a weight distribution of 0.5 for the main task and 0.1 for each of the auxiliary tasks. The error signals from auxiliary task group 1 were intended to be used to improve the generalization of the whole network, while those from auxiliary task group 2 were explicitly intended to improve the generalizability of the GRU layers. The other term is a penalizing term *P* introduced by Lin et al^10^.

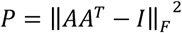

where *A* is the attention matrix whose rows are attention vectors ***a*** as introduced earlier, *I* is an identity matrix and ||·||*_F_* stands for the Frobenius norm of a matrix. This term encourages the diversity of the attention vectors ***a*** and is multiplied by a hyperparameter we can set arbitrarily.

### 2.5. Training

The models are trained with Adam optimizer with an early stopping rule requiring five consecutive failures to reduce the minimum loss of the main task. We applied multiplicative learning rate decay after 5^th^, 10^th^, 15^th^ and 30^th^ epoch with its decay rate parameterized for optimization. Supplementary Table 2 shows all the hyperparameters we used. They were optimized using tree-structured Parzen estimation with over 500 trials for each model^11^.

### 2.6. Performance evaluation

We assessed the performance of our AI systems measuring receiver operating characteristic (ROC) and precision-recall (PR) area under the curve (AUC) values. We used bootstrap resampling (N=2000) to calculate the 95% confidence intervals (CI) of the AUC values and to test the statistical significance of their differences. We set up a human expert vs. AI competition to evaluate the performance of our models. For this competition, we randomly sampled up to 10 cases without replacement from each outcome combination stratum (N=313), and then we added 200 additional cases sampled without stratification and replacement from the rest finalizing our final competition dataset (N total=513). The stratified sampling procedure was to increase the proportion of positive outcome cases, which will increase the statistical power of later comparison tests. A board-certified emergency medicine (EM) physician with 10-year experience as an EMS director predicted outcomes using the competition dataset. Using the results, we calculated recall (sensitivity) levels of the human expert and set the threshold levels of our models to achieve the same recall levels as the human expert. Lastly, we calculated and compared precision (positive predictive value, PPV), negative predictive value (NPV), and specificity levels of the models and the human expert. Their 95% confidence intervals and the significance of difference was assessed using bootstrap resampling.

### 2.7. Visualization and quality assessment of attention mapping

We visualize where our models are focusing on by drawing a heatmap over the tokens of the HPI sentences. The values of the heatmap are obtained by summing over all the attention vector ***a*** and rescaling the vector using min-max normalization (ranging 0 to 1).

A separate reviewer (a board-certified EM physician with two years of EMS director experience) rated the clinical relevance of the attention patterns in one hundred random samples from the test dataset using a 5-point Likert scale (Perfect, Good, Fair, Poor, Random). The reviewer was instructed to determine the quality based on general clinical relevance without being instructed for what purpose the models are used.

### 2.8. Statistical analysis

Categorical variables are reported using frequencies and proportions. Continuous variables are reported using the median and interquartile range (IQR). T-test, Wilcoxon’s rank-sum test, chi-square test, or Fisher’s exact test are performed as appropriate for comparison between groups.

P-values < 0.05 were considered significant. Neural network models were developed and tested using PyTorch package version 1.4 running on Python version 3.7^12^. Statistical analyses were performed on R-packages version 3.5.1 (R Foundation for Statistical Computing, Vienna, Austria).

## 3. Results

45,396 ED visits using the national EMS were identified. After the exclusion of OHCA, DOA, and transfer-out cases, 42,073 cases were included as the study population (Table 1). The median age of the population was 58.0 (43.0-73.0), and female cases were 21,023 (50.0%). The main outcome events including hospital admission, endotracheal intubation, mechanical ventilation, pressor infusion, surgery, cardiac catheterization, ICU admission and cardiac arrest occurred in 10,689 (25.4%), 915 (2.2%), 808 (1.9%), 1402 (3.3%), 1472 (3.5%), 783 (1.9%), 2226 (5.3%) and 310 (0.7%) cases, respectively. The number of cases in train, validation, and test dataset was 25,242, 8,414, and 8,414, respectively with no significant difference among the groups.

**Table 1.**
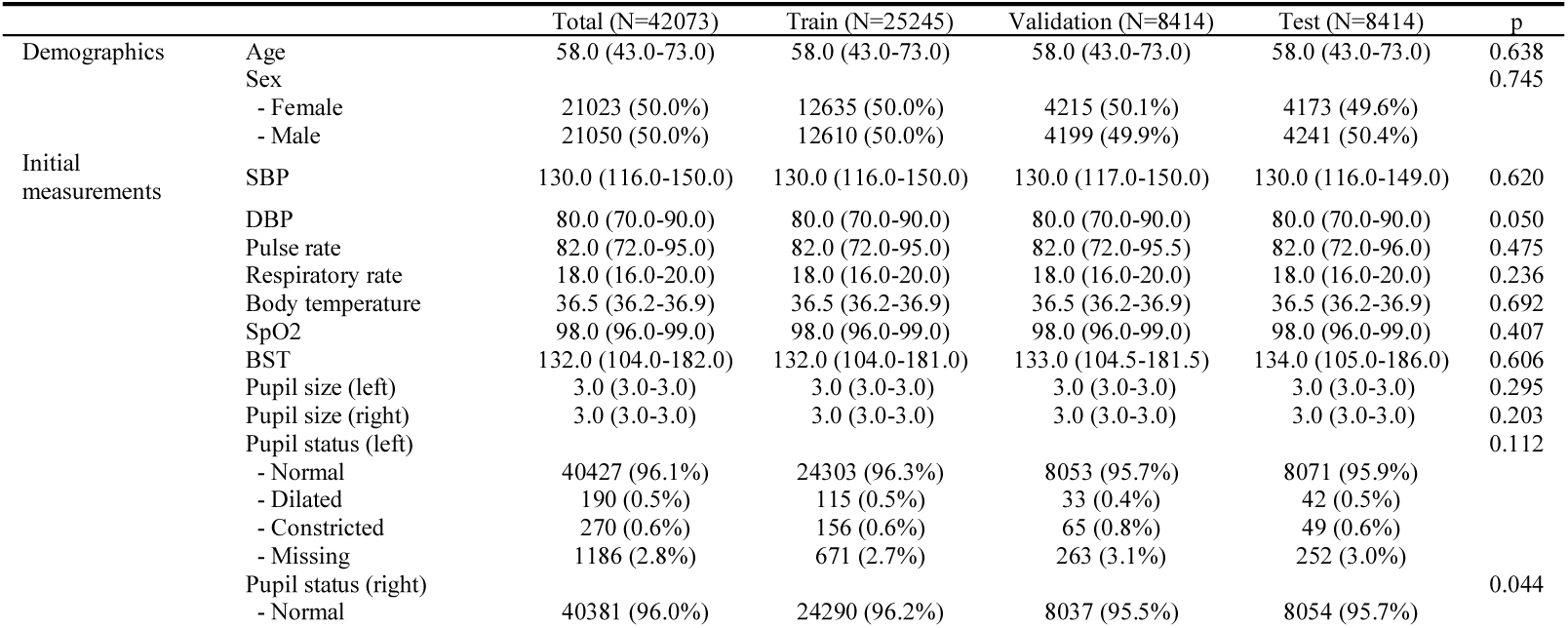
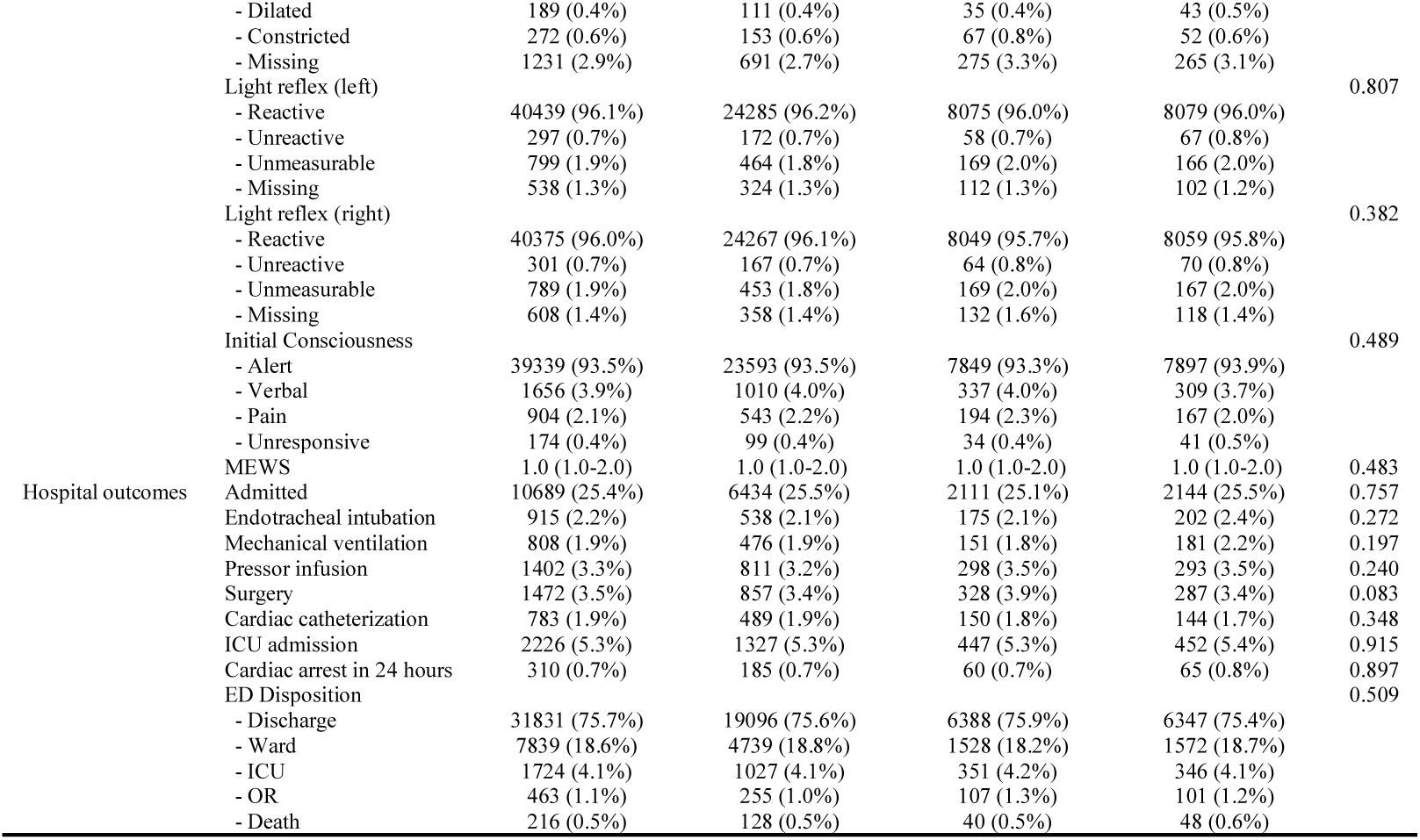
Study population

Figure 2 and 3 shows ROC and PR curves of the models, respectively, assessed in the test dataset. The ROC AUC values of the model I and P ranged from 0.793 to 0.929 and 0.812 to 0.934. respectively, both of which were higher to those of modified early warning score (MEWS) in every aspect (all p < 0.001, supplementary Table 3). PR AUC values ranged from 0.149 to 0.673 and 0.156 to 0.683, respectively, and were higher to those of MEWS (all p < 0.001). The model P generally performed better with significantly higher ROC AUC in the prediction of admission (p=0.017), mechanical ventilation (p=0.028), surgery (p=0.011), and cardiac arrest (p=0.006) and with significantly higher PR AUC in the prediction of admission (p<0.001), intubation (p=0.010), mechanical ventilation (p=0.005), and ICU admission (p=0.010).

**Fig. 2.**
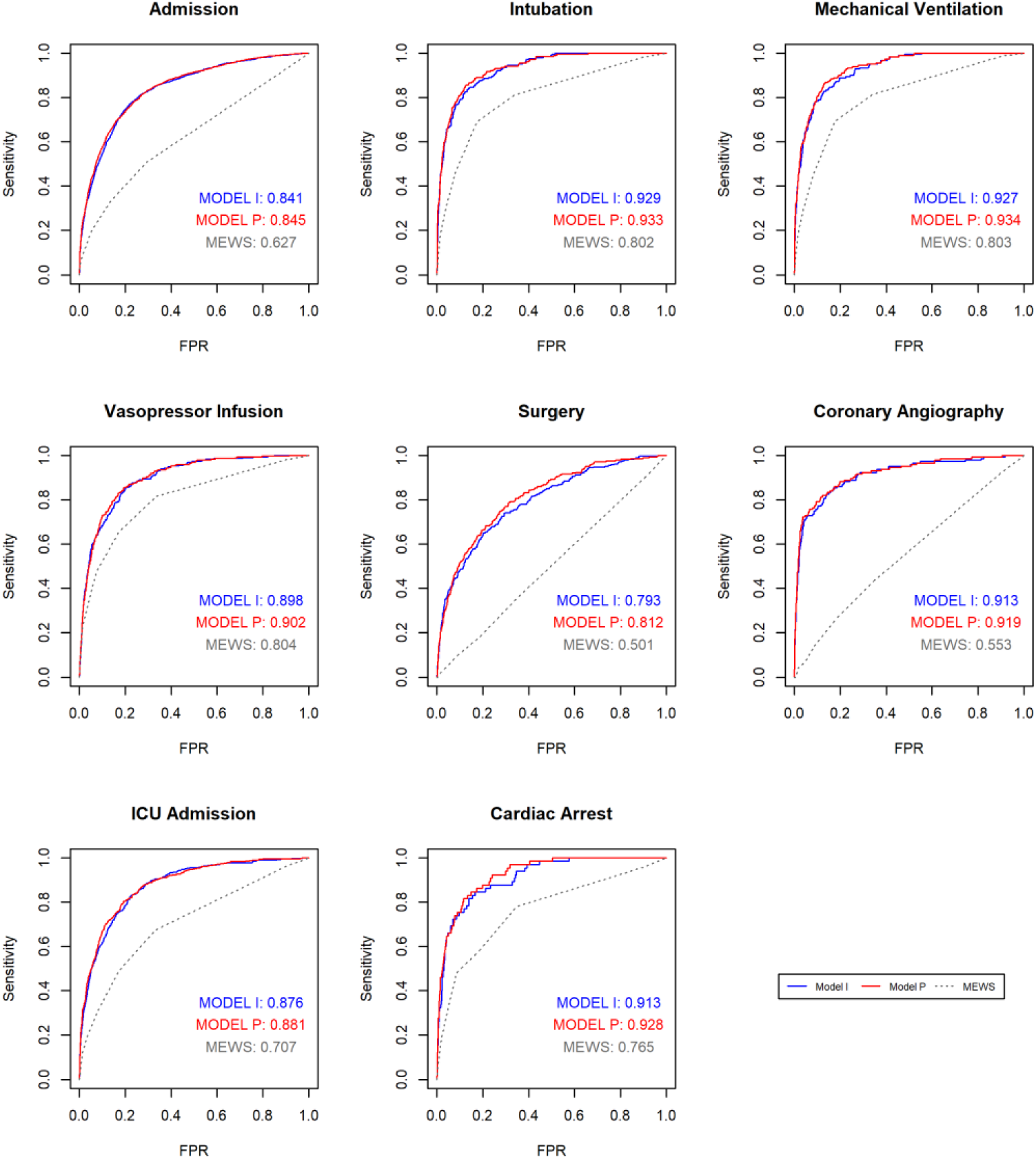
Receiver operating characteristic (ROC) curves of the model I (blue lines) and P (red lines) plotted against those of modified early warning score (MEWS, gray dashed lines) and their area under the curve (AUC) values; ICU, intensive care unit

**Fig. 3.**
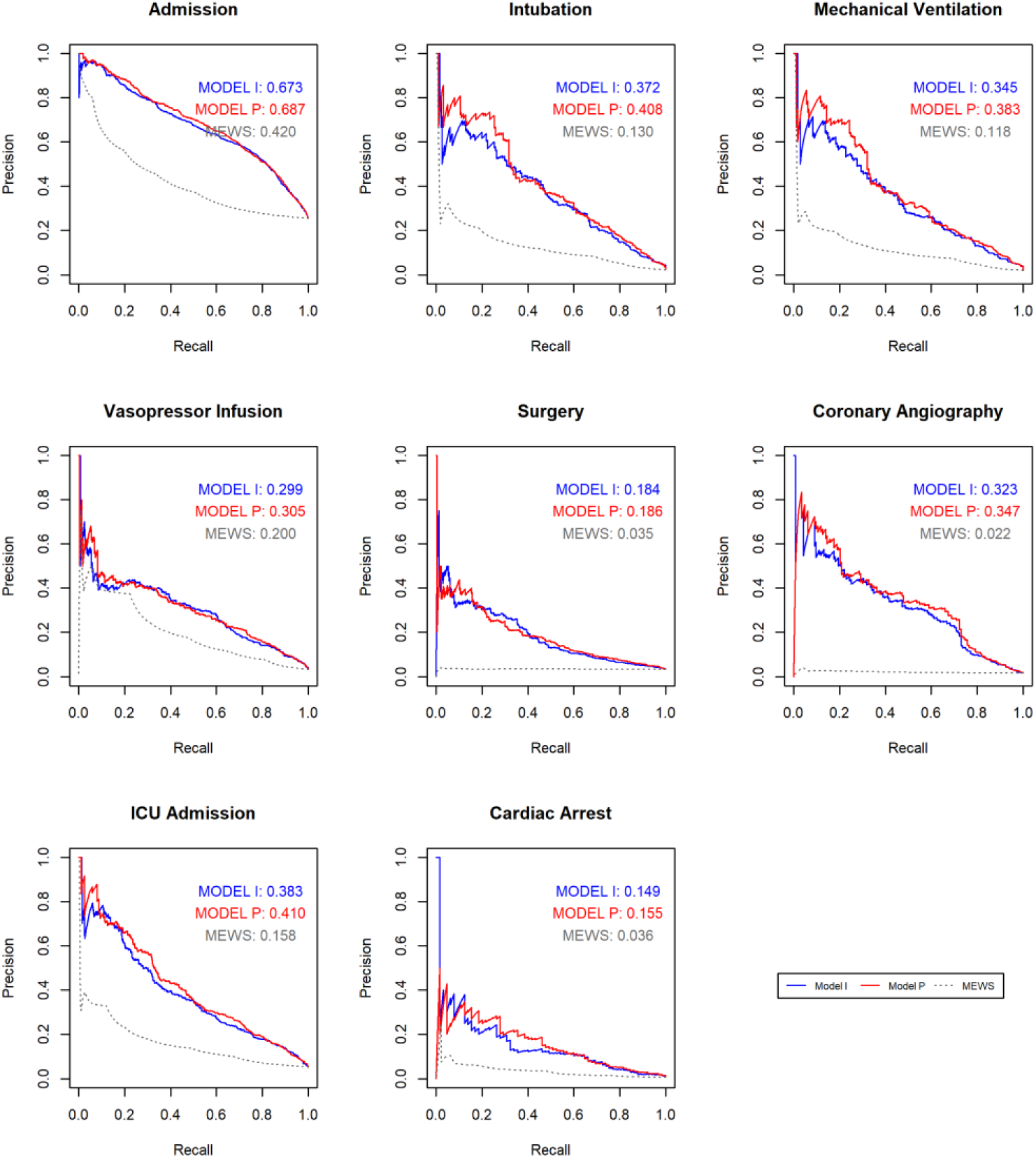
Precision-recall (PR) curves of the model I (blue lines) and P (red lines) plotted against those of modified early warning score (MEWS, gray dashed lines) and their area under the curve (AUC) values; ICU, intensive care unit

Figure 4 shows the results of a human expert vs. AI competition test. Our AI models achieved precision levels not significantly different from those of a human expert except in prediction of mechanical ventilation and ICU admission, where they achieved superior performance (p=0.030 [model I] and p=0.015 [model P], respectively, supplementary Table 4).

**Fig. 4.**
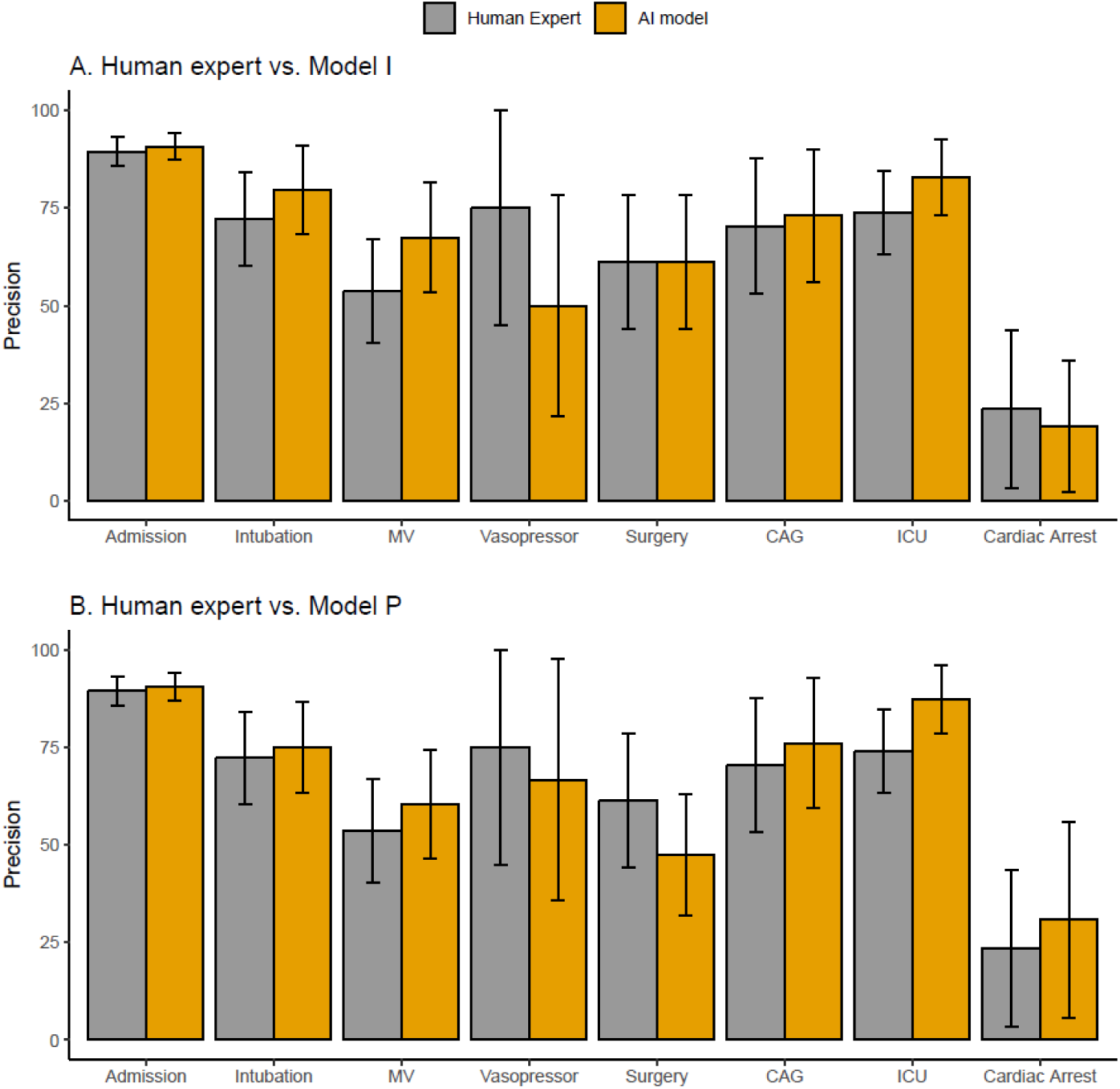
Precision comparison in the human expert vs. AI competition; MV, mechanical ventilation; CAG, coronary angiography; ICU, intensive care unit

Another human expert with two years of EMS director experience rated the quality of attention mappings in one hundred random samples from the test dataset (Figure 5). Only 10 percent of the cases were rated poor or worse (poor: 8, random: 2). Supplementary figure 1 shows the representative samples of attention maps chosen from the perfect/good/fair category cases.

**Fig. 5.**
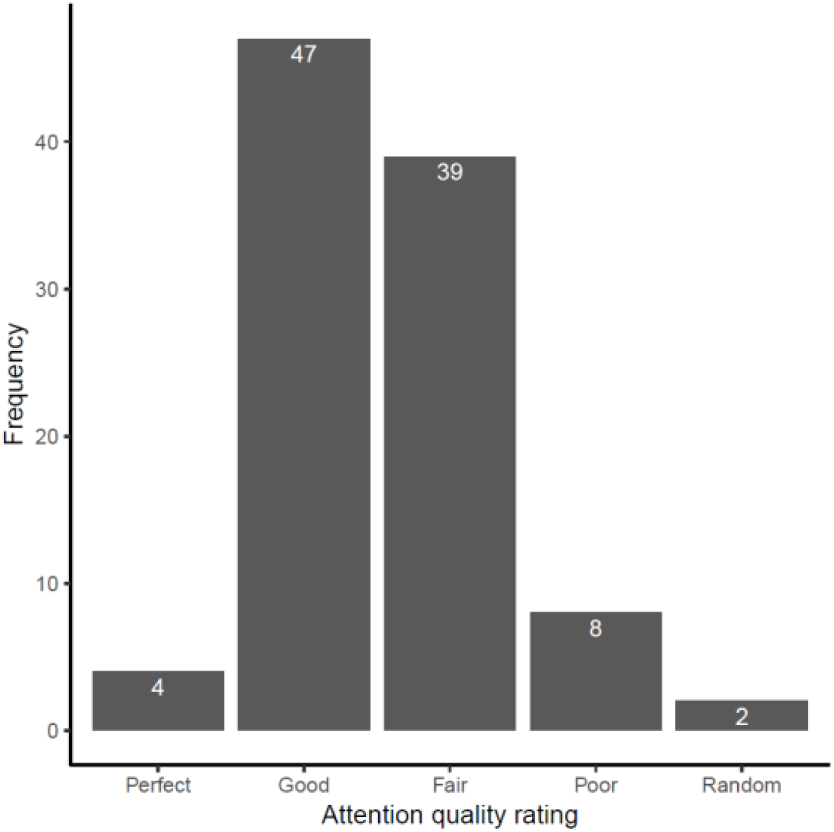
Quality ratings of attention mappings by a human expert in sample cases (N=100)

## 4. Discussion

In this study, we designed AI models that can jointly predict various hospital care needs using multimodal data at initial contact by EMS. The self-attention based-models were trained with multi-task learning methods. Our experiment showed that AI models could achieve similar or better performance than a human expert in this domain.

Accurate prediction of patients’ needs for hospital resources is critical because the type of hospitals and subsequent cares are dependent on it. ^2,13^Direct medical control can improve the quality of the prediction. However, it requires 24/7 access to EMS directors and may lead to increased workload. Several tools have been developed to help the decision process where a single yes or no type event (i.e., mortality or ICU admission) is predicted based on limited types and number of variables^14-16^. In this approach, one can achieve near-maximum performance allowed by the dataset using shallow algorithms (i.e., logistic regression with polynomial and interaction terms or other non-deep learning-based ML methods) if developed in a principled way^17,18^.

This approach, however, has obvious limitations. First, in many medical emergencies, a patient commonly has multiple care needs that cannot be predicted by a single output. Also, some of these predictions (i.e., surgery or coronary angiography) require target-specific features often scattered around in natural language data or in other unstructured data forms. In short, we need multiple outputs, each of which focusing on relevant information from both structured and unstructured data. This requirement motivated our adoption of self-attention mechanism. In the self-attention mechanism, the focus of attention is not hard-coded and determined by the input representation and queries. Through training, the mechanism learns to generate multiple attention patterns, each conditioned by each of the queries. The queries are also learnable and were parameterized by the W matrix in our models. How many queries a model needs for optimal performance would be task- and data-specific. The Bayesian hyperparameter optimization procedure suggested models with relatively many queries (eleven for both of the model types).

The size of our training dataset was relatively small. Considering the rare occurrence of the outcome events, it is surprising that our models achieved performance similar to a human expert. One of the possible reasons could be the use of multi-task learning. Health outcomes are often highly related to one another. One can take advantage of this by using multi-task learning, where a shared intermediate representation is used for multiple tasks. Possible benefits have been suggested^7^: 1) It has implicit data augmentation effect; 2) It helps the models to focus more on relevant features rather than noises; 3) It allows a task to use the features developed by another task; 4) It biases a model to prefer representations that other tasks also prefer which will help the model to be generalized to new tasks; 5) It acts as a regularizer by introducing an inductive bias which reduces the risk of overfitting. In our study, the feature-rich natural language input data could have made the models very prone to overfit. Our extensive application MTL may have provided significant benefits, probably in data augmentation and model regularization.

The model P performed better than model I. This suggests the contextual information should be repeatedly provided with each new input vector rather than being used once at the beginning of the unrolling of GRUs.

This could be explained by that the contextual information encoded in the initial hidden state will degrade as the unrolling progress and be “forgotten” eventually.

This study has several limitations. Firstly, the models were developed and tested using EMS records of a single hospital. The way of using natural language, as well as the population characteristics and clinical environment, can change by time and space. Therefore, we cannot guarantee the generalizability of our models. Second, only one human expert was compared to the AI models. We chose to use the performance scores of the most experienced EMS director as a comparator rather than averaged scores. Third, the role of direct medical control is much wider rather than some predictions of hospital care needs. Developing a system capable of providing all the expertise of an EMS director would be much more challenging.

Despite these limitations, our study has several strengths. This is the first study developing AI systems capable of jointly predicting multiple outcomes using only prehospital information. The system achieved human expert-level performance and provided interpretable outputs. Also, this is the first attempt to apply modern NLP techniques on EMS records. The models extract distributed sentence representations from unstructured real-world free text data were used for predictions. We also presented some clues on how contextual information can be incorporated into the computational graphs to improve prediction performance. Lastly, we also showed how multiple auxiliary tasks can be utilized in model development.

## 5. Conclusion

Our models with a self-attention mechanism trained using a multi-task learning method achieved similar (or superior) performance compared to an experienced human expert. Our study shows that AI models can be used to predict various medical resource requirements at initial contact by EMS.

## Data Availability

The research data is not open to the public.

---

**Supplementary Fig. 1.**
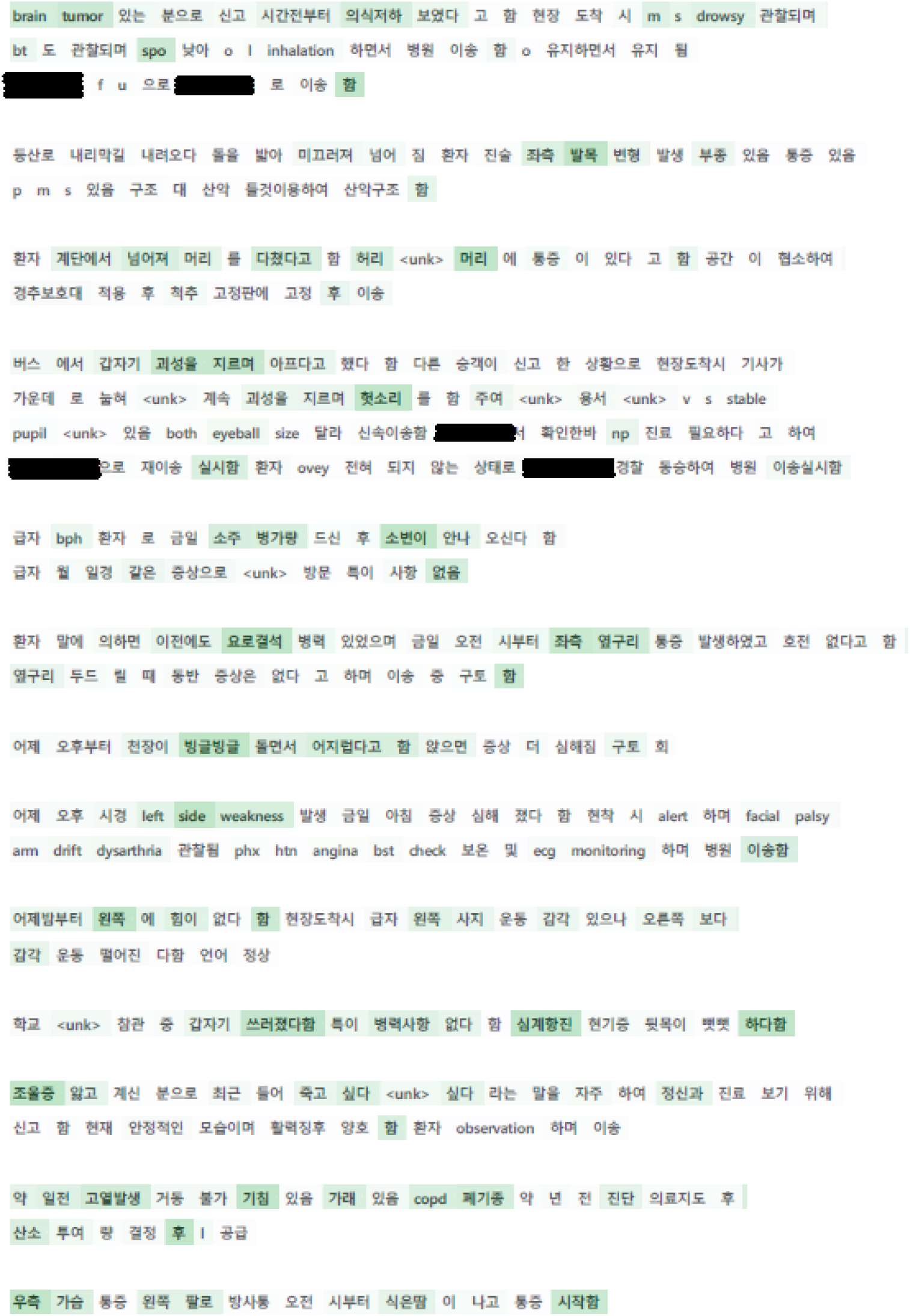
Examples of attention mappings generated by the models

## Acknowledgements

This work was supported by SNUBH grant No. 02-2013-060 and SNUBH grant No. 09-2015-001.

## Disclosures

The authors declare no conflict of interest.

## Notes

### Competing Interest Statement

The authors have declared no competing interest.

### Author Declarations

Seoul National University Bundang Hospital IRB

